# TP53 and Co-Occurring Pathogenic Variants Are Differentially Associated with Prognosis of Advanced Ovarian and Endometrial Carcinomas

**DOI:** 10.64898/2026.11.17.26368236

**Authors:** Janet Song, Shreyas Kudrimoti, Chen Jiang, Jenny Zhao, Yasmine Kamgarhaghighi, Hailey Liang, Ninah S. Achacoso, Aleyda V. Solorzano, Pamela Tse, Elaine Chung, Sachdev Thomas, Haruka Itakura, Talayeh Ghezelayagh, Laurel Habel, Minggui Pan

**Affiliations:** Internal Medicine Residency Program, Kaiser Permanente, Santa Clara, CA 94051, USA; Division of Research, Kaiser Permanente, Oakland, CA 94612, USA; Department of Oncology and Hematology, Kaiser Permanente, Vallejo, CA 94589, USA; Division of Oncology, Stanford University School of Medicine, Stanford, CA 94305, USA; Division of Gynecologic Oncology, Department of Obstetrics & Gynecology, Stanford University School of Medicine, Stanford, CA 94305, USA

**Keywords:** Ovarian carcinoma, Endometrial carcinoma, TP53, KRAS, Next-generation sequencing, Overall survival

## Abstract

**Objective:** To determine whether TP53 and other pathogenic variants (PVs) carry shared or organ-specific prognostic associations in advanced ovarian and endometrial carcinoma.

**Methods:** We analyzed 1060 patients with stage III–IV ovarian carcinoma (924 high-grade serous [HGSOC], 66 clear cell [CCOC], 42 endometrioid, 28 mucinous) and 543 with endometrial carcinoma (EC) profiled by next-generation sequencing. Associations with overall survival (OS) were estimated by multivariable Cox regression and combined across organs by fixed-effect pooling. Key findings were tested in 8881 tumors from public cBioPortal cohorts, including The Cancer Genome Atlas.

**Results:** TP53 PV was associated with worse OS in EC (HR 1.61, 95% CI 1.23–2.10) and CCOC (HR 3.23, 1.05–9.89) but not in HGSOC overall (HR 0.95, 0.72–1.25); in HGSOC, it was associated with worse OS only in KRAS-mutant tumors (HR 3.42, 1.13–10.33). KRAS PV was associated with better OS in CCOC (HR 0.29, 0.09–0.89) and HGSOC (HR 0.72, 0.50–1.02) but not in EC. In HGSOC, BRCA1/2 PV was associated with better OS and MYC amplification with worse OS. In EC, CTNNB1 and ARID1A PVs were associated with better OS. RB1 PV was associated with worse OS in both organs (pooled HR 1.47, 1.05–2.06). In public cohorts stratified by histology, TP53 alteration was associated with worse OS in EC (HR 1.69, 1.36– 2.10) and ovarian carcinoma (HR 1.28, 1.00–1.62).

**Conclusions:** The prognostic association of TP53 PV depends on organ, histotype and co- occurring KRAS PV, and RB1 PV is a shared adverse marker. These findings may inform prognostic stratification of Müllerian carcinomas.

## 1. Introduction

High-grade serous ovarian carcinoma (HGSOC) is the most common epithelial ovarian malignancy, followed by clear cell (CCOC), endometrioid (EOC) and mucinous (MOC) carcinoma; most patients present with advanced disease, and mortality remains high [1,2]. Endometrial carcinoma (EC) is the most common gynecologic malignancy, and patients with advanced or recurrent disease, particularly those with serous histology, have poor survival [3,4]. CCOC and MOC are often platinum-refractory, and effective treatment for these histotypes and for advanced EC remains a challenge [2,5,6].

BRCA1/2 PVs account for most hereditary HGSOC, and inhibitors of poly(ADP-ribose) polymerase (PARPi) are used to treat ovarian cancer with homologous recombination deficiency (HRD) [7–9]. In EC, molecular classification by POLE, mismatch repair and p53 status informs risk stratification, and immune checkpoint inhibitors have entered first-line treatment of advanced disease [10–14].

Ovarian and endometrial carcinomas arise from Müllerian epithelium and share recurrent alterations, including TP53, KRAS, PIK3CA, PTEN, ARID1A and CTNNB1 PVs and MYC amplification, although their distribution differs across histotypes [3,8,15]. TP53 PV is near- universal in HGSOC [16] and defines the copy-number-high, p53-abnormal class of EC, which carries the worst prognosis [10,11]. Whether TP53 PV, or any other shared alteration, carries the same prognostic weight at both organ sites, and whether co-occurring alterations modify it, remains unclear. In our previous studies, the prognostic associations of TP53 PVs in gastrointestinal adenocarcinomas depended on tumor context [17,18].

Here we analyzed 1603 patients with advanced ovarian carcinoma or EC profiled by tumor next- generation sequencing (NGS) within Kaiser Permanente Northern California (KPNC), aiming to identify PVs, TP53 in particular, whose prognostic associations are shared or organ-specific, and tested key findings in public cohorts.

## 2. Methods

### 2.1. Study population and data sources

KPNC is an integrated healthcare delivery system serving over 4 million members across 21 medical centers; since late 2017, NGS has been performed routinely for patients with advanced malignancy. The ovarian cohort included 1060 KPNC patients with stage III–IV HGSOC (n = 924), CCOC (n = 66), EOC (n = 42) or MOC (n = 28) whose tumors were profiled between November 2017 and December 2023. Histology was identified from pathology reports; primary sites included ovary, fallopian tube and peritoneum, and carcinosarcoma, germ cell, small cell, low-grade serous and benign histologies were excluded. The endometrial cohort included 543 patients with endometrioid (n = 257), serous (n = 224), mixed (n = 29) or other (n = 62) EC. Demographics, Charlson comorbidity index (CCI), Eastern Cooperative Oncology Group performance status (PS), stage, histology and systemic therapy were obtained from the electronic health record and cancer registry; CCI and race/ethnicity were not available for the endometrial cohort. The study was approved by the KPNC institutional review board with waiver of consent.

### 2.2. Sequencing and variant definitions

Tumors were profiled with StrataNGS, a 429-gene assay on co-isolated DNA and RNA from formalin-fixed, paraffin-embedded tissue [19]. In the ovarian cohort we analyzed the twelve most common alterations (TP53, BRCA1, BRCA2, KRAS, CDKN2A, PIK3CA, RB1, PTEN, BRAF, NF1 and ARID1A PVs and MYC amplification); in the endometrial cohort, the twenty most frequently altered genes. TP53 status was analyzed as any PV versus wild type (WT). For an exploratory analysis, TP53 PVs were further classified as hotspot gain-of-function (GOF; R175H, R248Q, R248W, R249S, R273H, R273L or R282W) or other [17]. HRD-associated genes included BRCA1, BRCA2, PALB2, ATM, CHEK1, CHEK2, Fanconi anemia pathway genes, BARD1, BRIP1, RAD51, ATR, ATRX, NBN, BLM and CDK12 [20]. PARPi (olaparib, rucaparib, niraparib) was given primarily as maintenance; all but seven ovarian patients (99.4%) received chemotherapy.

### 2.3. Statistical analysis

In the ovarian cohort, OS was measured from diagnosis of stage III–IV disease to death or end of follow-up (April 10, 2025; median follow-up 52.9 months); because NGS results became available a median of 7.7 months after diagnosis, Kaplan-Meier and Cox analyses accounted for delayed entry (left truncation) at the time of NGS [21]. In the endometrial cohort, OS was measured from date of diagnosis of advanced stage disease. Cox proportional hazards models estimated hazard ratios (HRs) and 95% confidence intervals (CIs). Ovarian models were adjusted for age, race/ethnicity, PS, CCI, histology and PARPi receipt; endometrial models were adjusted for age, PS, histology, and the multigene endometrial model included all twenty genes simultaneously. Effect modification of TP53 PV by co-occurring PVs was assessed within sub- cohorts and by product terms; co-alteration analyses compared four groups (both altered, each alone, neither). The proportional hazards assumption was assessed with Schoenfeld residuals [22]. For PVs analyzed in both cohorts, ovarian HRs were taken from a model including all twelve PVs in an analysis dataset of 1025 patients (Supplementary Table S1), and cohort- specific log HRs were combined by inverse-variance fixed-effect pooling with heterogeneity summarized by I² [23]. Because individual-level data were not merged, pooled estimates are exploratory. Analyses used SAS 9.4 and R.

### 2.4. External validation

Key findings were tested in public ovarian and endometrial cohorts accessed through cBioPortal [24], comprising 8881 tumors (3336 ovarian from 17 studies; 5545 endometrial from 22 studies), including The Cancer Genome Atlas (TCGA) PanCanAtlas, MSK-IMPACT, MSK-MET and CPTAC cohorts [25–27]. TP53 alteration was defined as a protein-altering mutation or deep deletion. OS was available for 2622 ovarian and 2985 endometrial tumors; HRs were estimated with Cox models unadjusted or stratified by histologic subtype

## 3. Results

### 3.1. Patient characteristics

Among 1603 patients, 1060 had ovarian carcinoma and 543 had EC (Table 1). Patients with CCOC, EOC and MOC were younger than those with HGSOC, and patients with CCOC were more often Asian (39.4% versus 16.8%) and less often received PARPi (7.6% versus 37.8%). Within EC, serous tumors occurred in older patients than endometrioid tumors (median 67.9 versus 64.9 years) and more often presented at stage IV (40.2% versus 28.8%).

**Table 1.** Clinical characteristics and frequency of PVs by organ and histology.

| Characteristic | HGSOC (n = 924) | CCOC (n = 66) | EOC (n = 42) | MOC (n = 28) | Endometrioid EC (n = 257) | Serous EC (n = 224) |
|---|---|---|---|---|---|---|
| *Clinical* |  |  |  |  |  |  |
| Age, median (range or IQR), years | 64 (26-94) | 55 (37-84) | NA | 54 (24-76)<sup>a</sup> | 64.9 (57.5-72.3) | 67.9 (61.8-73.0) |
| Asian race | 155 (16.8) | 26 (39.4) | NA | 5 (17.9)<sup>a</sup> | NA | NA |
| CCI, median (range) | 1 (0-11) | 0 (0-6) | NA | 0 (0-8)<sup>a</sup> | NA | NA |
| ECOG PS 0-1 | 687 (74.4) | 55 (83.3) | NA | 19 (67.9)<sup>a</sup> | 183 (71.2) | 171 (76.3) |
| Stage IV at initial diagnosis | —<sup>b</sup> | —<sup>b</sup> | —<sup>b</sup> | —<sup>b</sup> | 74 (28.8) | 90 (40.2) |
| PARPi received | 349 (37.8) | 5 (7.6) | NA | NA | NA | NA |
| Median OS, months | 33.4 | 33.8 | 66.8 | 25.1 | 31.0 | 23.1 |
| *Pathogenic variants* |  |  |  |  |  |  |
| TP53 PV | 762 (82.5) | 9 (13.6) | NA | 18 (64.3)<sup>a</sup> | 80 (31.1) | 197 (87.9) |
| BRCA1 PV | 99 (10.7) | 1 (1.5) | 2 (4.8) | 0 | —<sup>e</sup> | —<sup>e</sup> |
| BRCA2 PV | 52 (5.6) | 1 (1.5) | 3 (7.1) | 0 | —<sup>e</sup> | —<sup>e</sup> |
| KRAS PV | 74 (8.0) | 16 (24.2) | 13 (31.0) | 23 (82.1) | 64 (24.9) | 24 (10.7) |
| PIK3CA PV | 56 (6.1)<sup>c</sup> | 27 (40.9) | NA | 3 (10.7)<sup>a</sup> | 114 (44.4) | 80 (35.7) |
| PTEN PV | 22 (2.4)<sup>c</sup> | NA | NA | NA | 163 (63.4) | 27 (12.1) |
| ARID1A PV | 18 (1.9) | 20 (30.3) | NA | 2 (7.1)<sup>a</sup> | 51 (19.8) | 7 (3.1) |
| CTNNB1 PV | —<sup>e</sup> | —<sup>e</sup> | —<sup>e</sup> | —<sup>e</sup> | 65 (25.3) | 5 (2.2) |
| MYC amplification | 59 (6.4) | 0 | 3 (7.1) | 0 | 15 (5.8) | 11 (5.6)<sup>d</sup> |
| RB1 PV | 28 (3.0)<sup>c</sup> | NA | NA | NA | 15 (5.8) | 10 (5.1)<sup>d</sup> |
| CDKN2A PV | 26 (2.8)<sup>c</sup> | NA | NA | NA | 11 (4.3) | 8 (4.1)<sup>d</sup> |
| CCNE1 amplification | 30 (3.3)<sup>c</sup> | NA | NA | NA | 5 (1.9) | 11 (5.6)<sup>d</sup> |
| NF1 PV | 27 (2.9)<sup>c</sup> | NA | NA | NA | —<sup>e</sup> | —<sup>e</sup> |

Values are No. (%) unless noted. Age is median (range) for ovarian and median (IQR) for endometrial carcinoma. Ovarian values are from the KPNC ovarian cohort. <sup>a</sup>From the ovarian analysis dataset (n = 1025). <sup>b</sup>All ovarian patients had stage 3 or 4 disease at cohort entry. <sup>c</sup>Denominator 922 HGSOC with BRCA1/2 status. <sup>d</sup>Denominator 195 serous EC in the endometrial mutation table. <sup>e</sup>Not among the PVs analyzed in that cohort. NA, not available in the source data [obtain EOC demographics and the remaining NA cells from the KPNC analysis dataset]. Abbreviations: CCI, Charlson comorbidity index; CCOC, clear cell ovarian carcinoma; EC, endometrial carcinoma; EOC, endometrioid ovarian carcinoma; HGSOC, high-grade serous ovarian carcinoma; IQR, interquartile range; MOC, mucinous ovarian carcinoma; OS, overall survival; PARPi, poly(ADP-ribose) polymerase inhibitor; PS, performance status; PV, pathogenic variant.

PV distribution differed by organ and histotype (Table 1, Supplementary Fig. S1). TP53 PV was present in 82.5% of HGSOC, 64.3% of MOC, 13.6% of CCOC, 87.9% of serous EC and 31.1% of endometrioid EC. KRAS PV was present in 82.1% of MOC, 31.0% of EOC, 24.2% of CCOC and 8.0% of HGSOC, and in 24.9% of endometrioid and 10.7% of serous EC. PTEN (63.4%), PIK3CA (44.4%), CTNNB1 (25.3%) and ARID1A (19.8%) PVs predominated in endometrioid EC, and PIK3CA (40.9%) and ARID1A (30.3%) in CCOC. MYC amplification was present in 6.4% of HGSOC and 6.1% of EC; in HGSOC it occurred in 12.4% of BRCA1 PV, none of BRCA2 PV and 6.1% of BRCA1/2 WT tumors, while KRAS PV occurred in none of BRCA1 PV and 9.3% of BRCA1/2 WT tumors (Supplementary Table S3).

### 3.2. Survival by histology

Median OS was 33.4, 33.8, 66.8 and 25.1 months for HGSOC, CCOC, EOC and MOC (Fig. 1A). Compared with HGSOC, OS was worse for MOC (HR 2.10, 1.23–3.59), similar for CCOC (HR 1.04, 0.68–1.60) and better for EOC (HR 0.62, 0.36–1.07) (Fig. 1B). In EC, median OS was 27.2 months and was worse for serous than endometrioid tumors (23.1 versus 31.0 months, p < 0.001) (Fig. 1C).

**Fig. 1.**
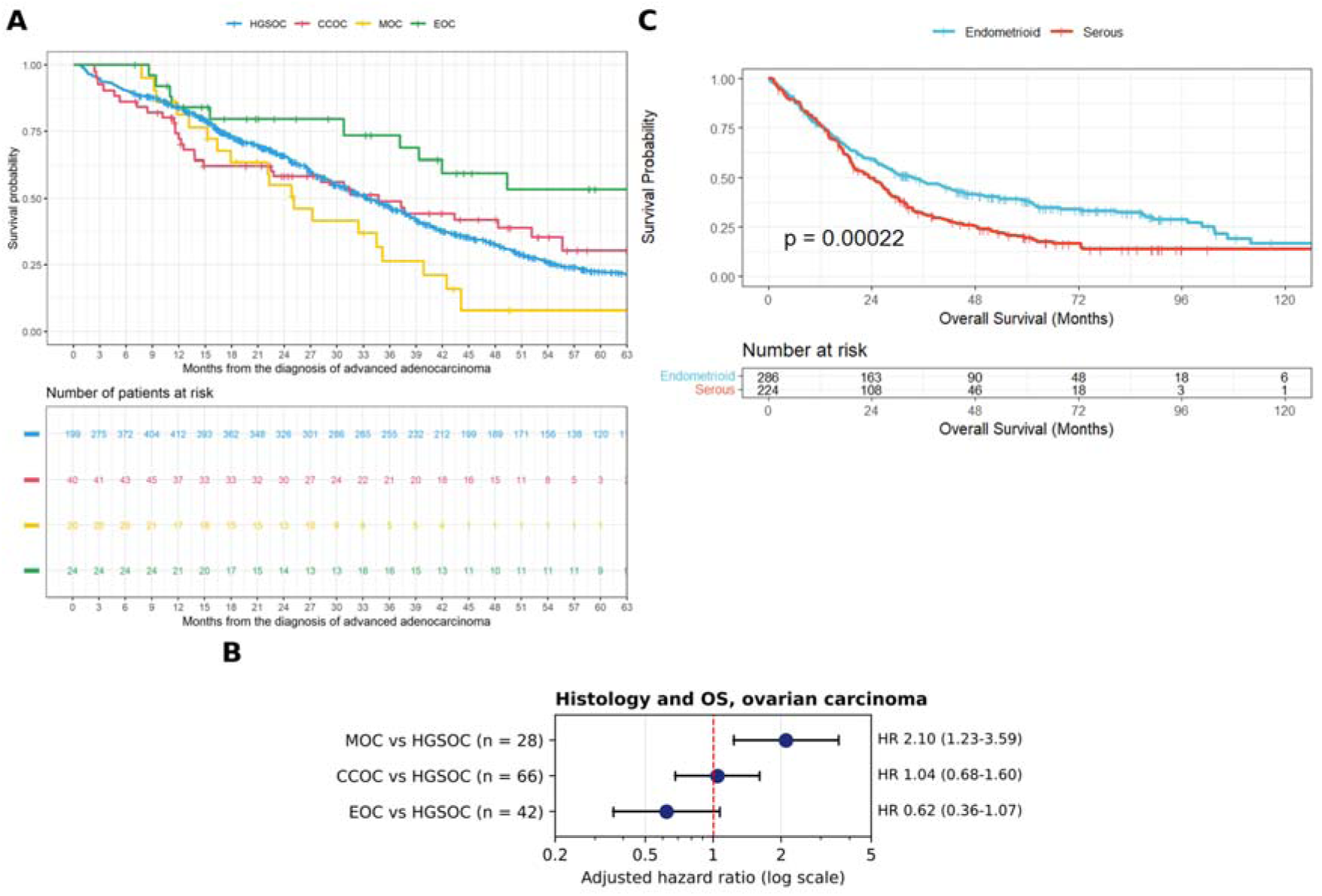
Survival by histology in ovarian and endometrial carcinoma. (A) Kaplan-Meier OS by ovarian histotype; median OS 33.4 (HGSOC), 33.8 (CCOC), 25.1 (MOC) and 66.8 (EOC) months. Numbers at risk account for delayed entry at the time of NGS. (B) Adjusted HRs for OS of MOC, CCOC and EOC versus HGSOC. (C) Kaplan-Meier OS of serous versus endometrioid EC (log-rank p < 0.001); median OS 23.1 and 31.0 months.

### 3.3. PVs and OS in ovarian carcinoma

TP53 PV was associated with worse OS in CCOC (HR 3.23, 1.05–9.89; 7 of 9 versus 25 of 57 patients died) but not in HGSOC (HR 0.95, 0.72–1.25) or in all ovarian carcinoma in the twelve- PV model (HR 1.14, 0.89–1.47) (Fig. 2A, Supplementary Fig. S4). In HGSOC, the TP53 association depended on KRAS status: within KRAS-mutant tumors, TP53 PV was associated with worse OS (HR 3.42, 1.13–10.33; 28 of 38 versus 12 of 36 died), whereas within KRAS-WT tumors it was not (HR 0.95, 0.72–1.25; interaction HR 2.06, 0.98–4.35) (Fig. 2A–C). Results were similar in the analysis dataset including all histologies (KRAS-mutant HR 2.13, 1.13–3.99; KRAS-WT HR 0.99, 0.77–1.27), and TP53 PV was not associated with OS within BRCA1/2- mutant or PIK3CA-mutant tumors (Supplementary Table S2).

**Fig. 2.**
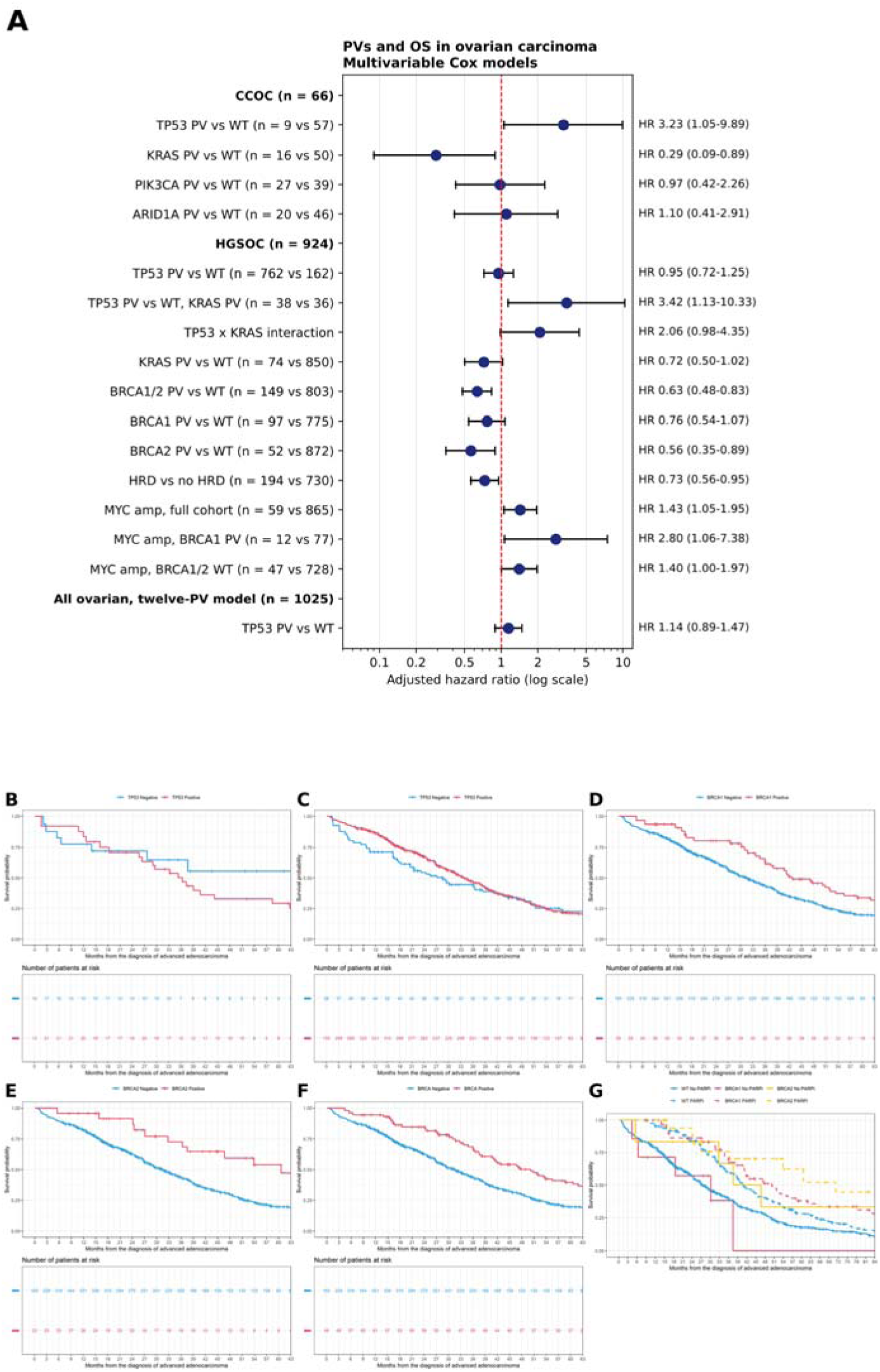
PVs and OS in ovarian carcinoma. (A) Adjusted HRs for OS in CCOC, HGSOC and all ovarian carcinoma (twelve-PV model), with numbers of patients. (B, C) Kaplan-Meier OS of TP53 PV versus WT within KRAS-mutant (p = 0.029) and KRAS-WT (p = 0.712) HGSOC. (D–F) Kaplan-Meier OS of HGSOC with BRCA1 PV (p = 0.030), BRCA2 PV (p = 0.019) and BRCA1/2 PV combined (p = 0.002) versus WT. (G) Kaplan-Meier OS by PARPi receipt within BRCA1 PV, BRCA2 PV and BRCA1/2 WT HGSOC.

KRAS PV was associated with better OS in CCOC (HR 0.29, 0.09–0.89; 6 of 16 versus 26 of 50 died) and HGSOC (HR 0.72, 0.50–1.02), with similar results in BRCA1/2 WT HGSOC (HR 0.70, 0.48–1.00); PIK3CA and ARID1A PVs were not associated with OS in CCOC (Fig. 2A).

In HGSOC, BRCA1/2 PV was associated with better OS (HR 0.63, 0.48–0.83), driven mainly by BRCA2 PV (HR 0.56, 0.35–0.89; BRCA1 PV HR 0.76, 0.54–1.07), as was HRD (HR 0.73, 0.56–0.95) (Fig. 2A). Median OS was 42.7 months for BRCA1 PV, 60.6 months for BRCA2 PV and 31.6 months for BRCA1/2 WT (Fig. 2D–F). PARPi receipt was associated with longer OS in each BRCA group: 50.1 versus 30.2 months for BRCA1 PV, 71.2 versus 40.1 months for BRCA2 PV and 40.9 versus 27.7 months for BRCA1/2 WT (Fig. 2G). MYC amplification was associated with worse OS (HR 1.43, 1.05–1.95; 46 of 59 versus 435 of 865 died), including within BRCA1 PV (HR 2.80, 1.06–7.38) and BRCA1/2 WT (HR 1.40, 1.00–1.97) tumors. In the twelve-PV model, RB1 PV was associated with worse OS (HR 1.79, 1.13–2.84), with trends for NF1 (HR 1.70, 0.97–2.97) and CDKN2A (HR 1.38, 0.90–2.10) (Supplementary Table S1,

Supplementary Fig. S3).

### 3.4. PVs and OS in endometrial carcinoma

TP53 PV, present in 314 patients (57.8%), was associated with worse OS (n = 304 versus 206 with follow-up; log-rank p < 0.0001) (Fig. 3B). Adjusted for age and PS, TP53 PV was associated with worse OS irrespective of co-occurring KRAS PV (TP53 alone HR 1.61, 1.23– 2.10; both HR 1.46, 0.95–2.24; KRAS alone HR 0.94, 0.62–1.43) or PIK3CA PV (TP53 alone HR 1.55, 1.14–2.10; both HR 1.54, 1.09–2.16; PIK3CA alone HR 0.91, 0.63–1.32), relative to neither alteration (Fig. 3D, E). The TP53 association was attenuated in the twenty-gene model (HR 1.24, 0.94–1.64) (Fig. 3A), consistent with the concentration of TP53 PV in serous tumors.

**Fig. 3.**
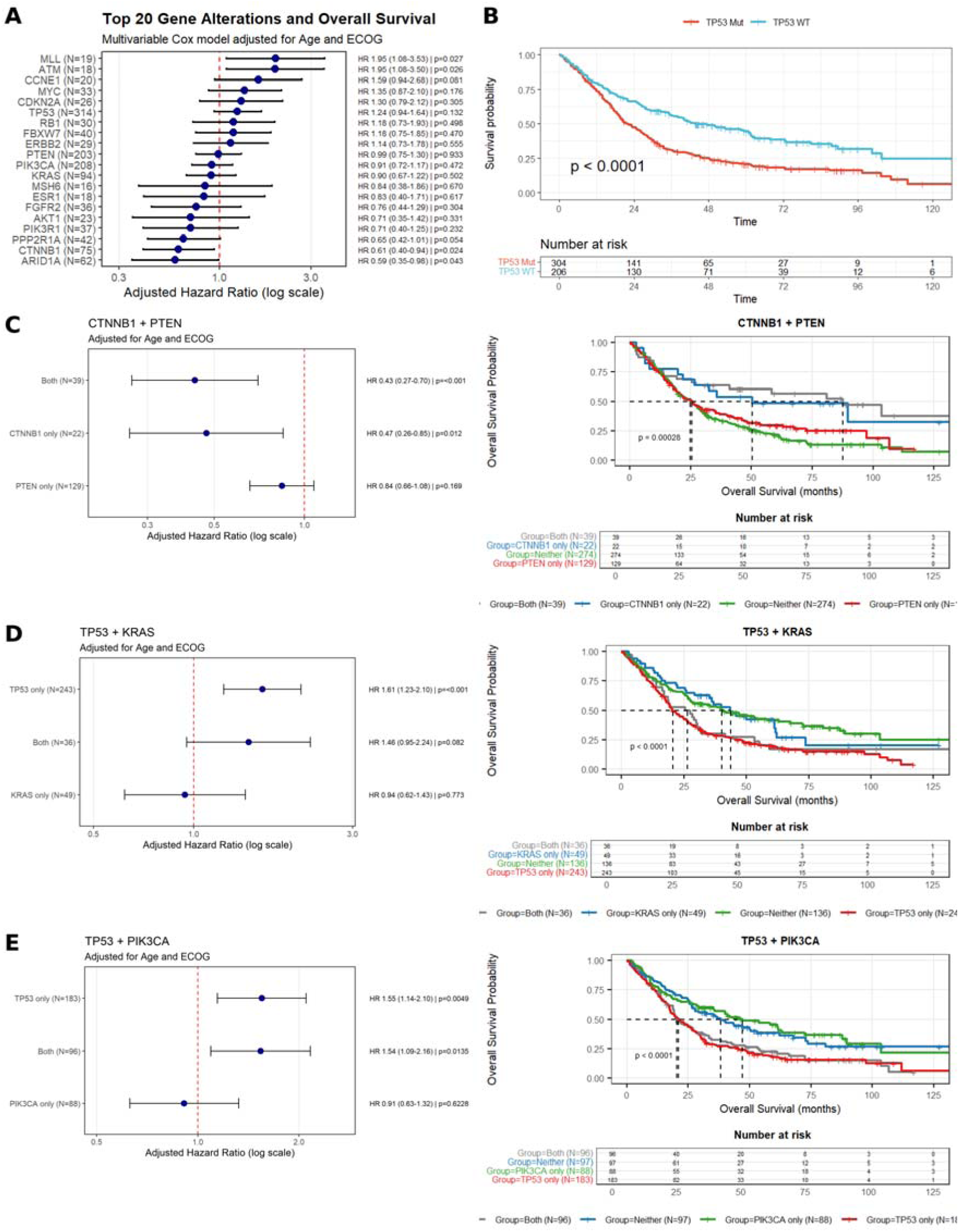
PVs and OS in endometrial carcinoma. (A) HRs for the twenty most common PVs in a multigene model adjusted for age and PS. (B) Kaplan-Meier OS of TP53 PV (n = 304) versus WT (n = 206) (log-rank p < 0.0001). (C–E) HRs and Kaplan-Meier OS for CTNNB1 and PTEN (C), TP53 and KRAS (D), and TP53 and PIK3CA (E) co-alteration groups, relative to neither alteration and adjusted for age and PS.

In the twenty-gene model, CTNNB1 (HR 0.61, 0.40–0.94) and ARID1A (HR 0.59, 0.35–0.98) PVs were associated with better OS, and MLL (HR 1.95, 1.08–3.53) and ATM (HR 1.95, 1.08– 3.50) PVs with worse OS; CCNE1 amplification (HR 1.59, 0.94–2.68) and PPP2R1A PV (HR 0.65, 0.42–1.01) showed borderline associations (Fig. 3A). Co-occurring CTNNB1 and PTEN PVs (n = 39; HR 0.43, 0.27–0.70) and CTNNB1 PV alone (n = 22; HR 0.47, 0.26–0.85) were associated with better OS than neither, whereas PTEN PV alone was not (HR 0.84, 0.66–1.08); median OS exceeded 85 months with both alterations (Fig. 3C). PTEN and PIK3CA co- alteration groups did not differ significantly (Supplementary Fig. S2).

### 3.5. Shared and organ-specific associations

Among PVs analyzed in both cohorts (Table 2, Supplementary Fig. S5), TP53 PV had directionally concordant multivariable HRs in ovarian carcinoma (HR 1.14) and EC (HR 1.24) (pooled HR 1.18, 0.98–1.43; I² 0%), but its association varied by histotype and co-alteration, being adverse in EC, CCOC and KRAS-mutant HGSOC and null in HGSOC overall. RB1 PV was associated with worse OS at both sites (ovarian HR 1.79, EC HR 1.18; pooled HR 1.47, 1.05–2.06; I² 34%), and MYC amplification (pooled HR 1.36, 1.05–1.75; I² 0%) and CDKN2A PV (pooled HR 1.34, 0.97–1.85; I² 0%) showed concordant adverse associations. KRAS PV was associated with better OS in ovarian carcinoma (HR 0.71, 0.52–0.98) but not in EC. ARID1A PV (I² 59%) and CCNE1 amplification (I² 81%) showed opposite directions in the two organs, and PIK3CA and PTEN PVs were not associated with OS in either. Classifying TP53 PVs as hotspot GOF or other did not distinguish OS in HGSOC (HR 1.06, 0.80–1.38) or EC (HR 1.12, 0.81– 1.55) (Supplementary Table S5, Supplementary Fig. S6).

**Table 2.**
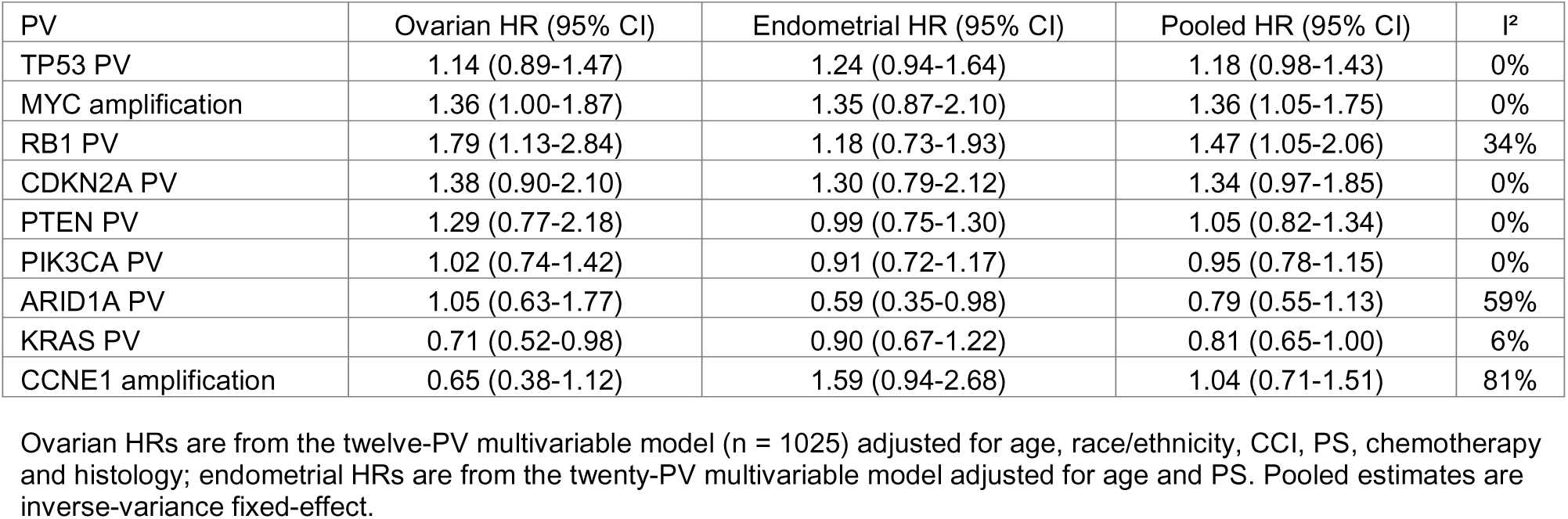
Multivariable HRs for OS of PVs analyzed in both cohorts.

| PV | Ovarian HR (95% CI) | Endometrial HR (95% CI) | Pooled HR (95% CI) | I<sup>2</sup> |
|---|---|---|---|---|
| TP53 PV | 1.14 (0.89-1.47) | 1.24 (0.94-1.64) | 1.18 (0.98-1.43) | 0% |
| MYC amplification | 1.36 (1.00-1.87) | 1.35 (0.87-2.10) | 1.36 (1.05-1.75) | 0% |
| RB1 PV | 1.79 (1.13-2.84) | 1.18 (0.73-1.93) | 1.47 (1.05-2.06) | 34% |
| CDKN2A PV | 1.38 (0.90-2.10) | 1.30 (0.79-2.12) | 1.34 (0.97-1.85) | 0% |
| PTEN PV | 1.29 (0.77-2.18) | 0.99 (0.75-1.30) | 1.05 (0.82-1.34) | 0% |
| PIK3CA PV | 1.02 (0.74-1.42) | 0.91 (0.72-1.17) | 0.95 (0.78-1.15) | 0% |
| ARID1A PV | 1.05 (0.63-1.77) | 0.59 (0.35-0.98) | 0.79 (0.55-1.13) | 59% |
| KRAS PV | 0.71 (0.52-0.98) | 0.90 (0.67-1.22) | 0.81 (0.65-1.00) | 6% |
| CCNE1 amplification | 0.65 (0.38-1.12) | 1.59 (0.94-2.68) | 1.04 (0.71-1.51) | 81% |

Ovarian HRs are from the twelve-PV multivariable model (n = 1025) adjusted for age, race/ethnicity, CCI, PS, chemotherapy and histology; endometrial HRs are from the twenty-PV multivariable model adjusted for age and PS. Pooled estimates are inverse-variance fixed-effect.

### 3.6. Validation in public cohorts

TP53 alteration prevalence in the public cohort was similar to KPNC in CCOC (11.2% versus 13.6%), MOC (74.5% versus 64.3%) and serous EC (95.2% versus 87.9%), higher in HGSOC (95.1% versus 82.5%) and lower in endometrioid EC (19.0% versus 31.1%), which in the public cohort included early-stage disease (Fig. 4A, Supplementary Table S4). TP53 alteration was associated with worse OS in public EC (unadjusted HR 2.58, 2.22–2.99) and ovarian carcinoma (HR 1.50, 1.27–1.77). Among tumors without KRAS PV and stratified by histology, the EC estimate (HR 1.69, 1.36–2.10) closely matched that in KPNC (HR 1.61, 1.23–2.10), and the ovarian estimate was attenuated (HR 1.28, 1.00–1.62) (Fig. 4C). The TP53 × KRAS interaction was directionally replicated in ovarian carcinoma (HR 1.87, 0.97–3.62) but was inverse in EC (HR 0.63, 0.42–0.95). KRAS PV without TP53 alteration was associated with better OS in public ovarian carcinoma (unadjusted HR 0.64, 0.42–0.97; histology-stratified HR 0.70, 0.43–1.14) but not in EC (stratified HR 1.24, 0.95–1.62). MYC amplification showed a weaker association in public HGSOC (HR 1.08, 0.83–1.41; n = 1427) than in KPNC; the difference was not significant (p = 0.21), and the pooled HR was 1.22 (1.01–1.46).

**Fig. 4.**
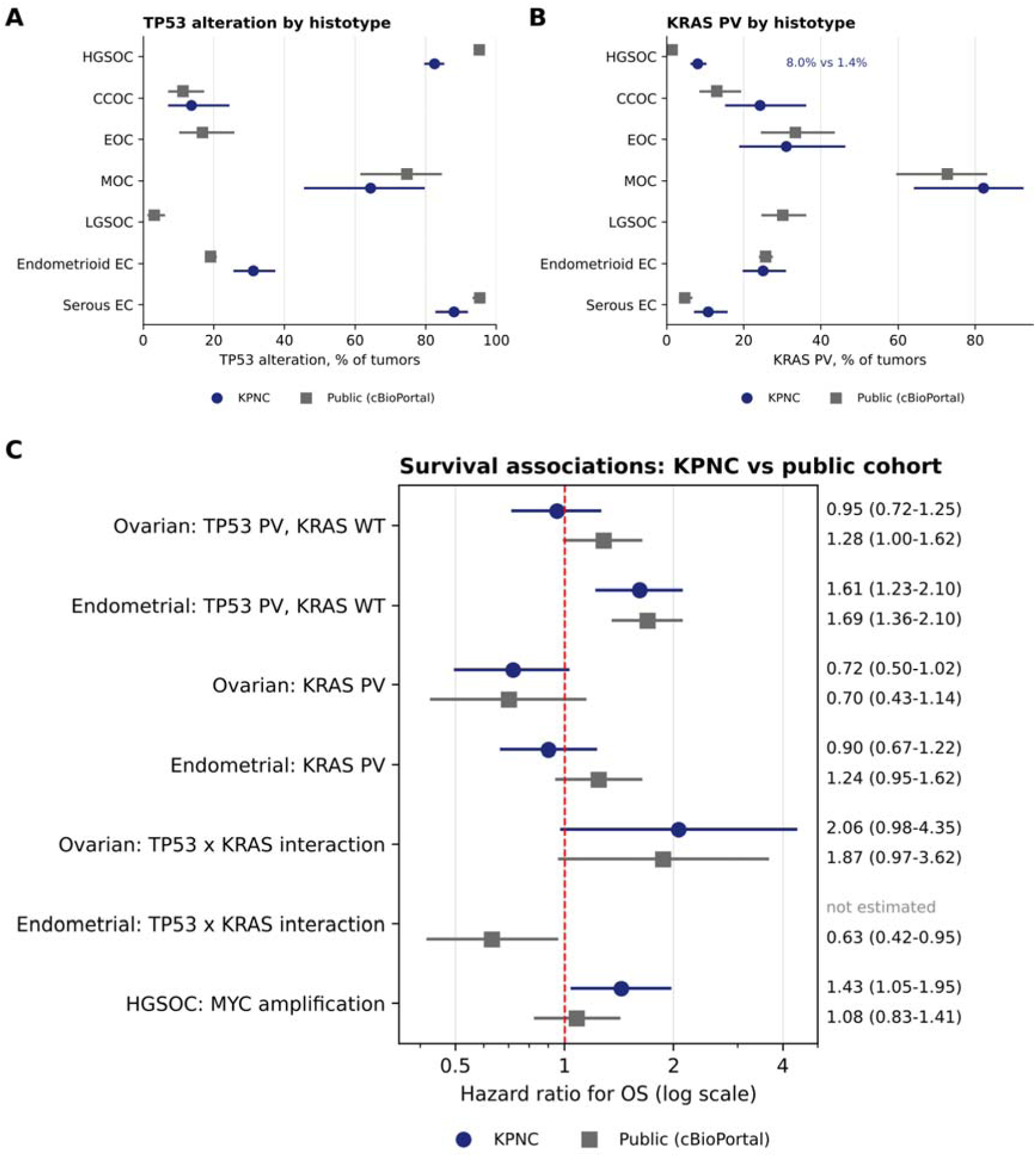
Validation in public ovarian and endometrial cohorts (cBioPortal, including TCGA). (A) TP53 alteration and (B) KRAS PV prevalence by histotype with 95% CIs; low-grade serous ovarian carcinoma (LGSOC) was excluded from the KPNC cohort, and TP53 status for KPNC EOC was not available. (C) HRs for OS in the KPNC cohort (multivariable; ovarian TP53 estimate from HGSOC) and the public cohort (stratified by histology, or unadjusted for the interaction and MYC estimates). TP53 PV, KRAS WT denotes TP53 alteration without KRAS PV versus neither.

KRAS PV was present in 1.4% of public HGSOC versus 8.0% of KPNC HGSOC, and in 30.1% of public low-grade serous carcinoma (Fig. 4B). Using public reference rates, the TP53 deficit and KRAS excess each implied that approximately 14% and 23% of KPNC HGSOC had a non- HGSOC molecular profile. KRAS prevalence in CCOC, EOC, MOC and endometrioid EC was similar between cohorts.

## 4. Discussion

In 1603 patients with advanced ovarian carcinoma or EC, the prognostic association of TP53 PV depended on organ, histotype and co-occurring KRAS PV. TP53 PV was associated with worse OS in EC and CCOC, and in HGSOC only when KRAS PV co-occurred, and public cohorts confirmed its adverse association in both organs. RB1 PV was associated with worse OS at both organ sites and MYC amplification showed a concordant but more modest association, whereas KRAS PV was associated with better OS in ovarian carcinoma but not in EC, and CTNNB1 and ARID1A PVs with better OS in EC. Improved OS with BRCA1/2 PV, particularly BRCA2, and with PARPi is consistent with prior reports [28–30].

In EC, TP53 PV marks the copy-number-high, p53-abnormal molecular class, which carries the worst prognosis across histologies [10,11,31]. Its adverse association in our advanced-stage cohort persisted irrespective of co-occurring KRAS or PIK3CA PV and was replicated after stratification by histology in nearly 3000 public tumors, indicating that TP53 status carries prognostic information beyond histotype; its attenuation in the twenty-gene model likely reflects overlap with serous histology and other copy-number-high alterations. In HGSOC, in which TP53 PV is near-universal [16], TP53 status was not prognostic overall, consistent with prior series [8,32], and the few TP53-WT tumors are likely heterogeneous. TP53 PV was associated with worse OS in CCOC, consistent with a previous report [33], where it may identify a genomically unstable, high-grade subset. In our previous studies, the prognostic associations of TP53 PVs depended on tumor context [17,18]; in Müllerian carcinomas, distinguishing hotspot GOF from other TP53 PVs added no prognostic information, supporting TP53 PV status as a whole as the clinically relevant variable.

The adverse association of TP53 PV within KRAS-mutant HGSOC, directionally replicated in public ovarian data, suggests that KRAS-mutant ovarian tumors without TP53 PV carry favorable prognosis, whereas co-occurring TP53 PV marks high-grade biology. Because TP53 PVs are near-universal in HGSOC [16], TP53-WT, KRAS-mutant “HGSOC” may in part represent misclassified low-grade serous carcinoma; the public comparison supports this concern, implying that 14–23% of KPNC HGSOC had a non-HGSOC profile. Nevertheless, the favorable KRAS association was replicated in public ovarian carcinoma, persisted in attenuated form after stratification by histology, and was present in CCOC. KRAS PV is generally associated with worse OS in other malignancies [17,34]; its favorable association in ovarian but not endometrial carcinoma, and the inverse TP53 × KRAS interaction in public EC, suggest biology specific to ovarian disease or its histotype composition rather than a general property of KRAS in Müllerian epithelium. Central pathology review of KRAS-mutant HGSOC is warranted.

RB1 PV was associated with worse OS in ovarian carcinoma, and the concordant direction in EC, together with a similar trend for CDKN2A PV, supports disruption of the CDKN2A–RB1 axis as a shared adverse feature of Müllerian carcinomas. MYC amplification was associated with worse OS in KPNC HGSOC, including BRCA1 PV tumors, consistent with its adverse role in lymphoma and osteosarcoma [35,36], but the association was weaker in public HGSOC; differences in amplification calling, suggested by the lower MYC amplification frequency in KPNC, and in covariate adjustment may contribute. CCNE1 amplification and ARID1A PV showed opposite directions in the two organs and warrant confirmation.

In EC, CTNNB1 and ARID1A PVs were associated with better OS, and co-occurring CTNNB1 and PTEN PVs identified a subgroup with median OS exceeding 85 months. CTNNB1 PV has been associated with recurrence in early-stage, low-grade endometrioid EC [37]; in our cohort of advanced or recurrent disease, it instead marked a favorable subgroup, likely reflecting enrichment in copy-number-low endometrioid tumors.

Strengths of this study include more than 1600 patients profiled by comprehensive NGS in an integrated health system, adjustment for PARPi in the ovarian cohort, reporting of events for each comparison, analysis of TP53 within co-alteration strata, and validation in more than 8800 public tumors. Limitations include the retrospective design and delayed NGS testing, addressed by left-truncation methods [21]; individual-level data were not merged; endometrial models were not adjusted for histology, molecular class, CCI, race/ethnicity or treatment, including immune checkpoint inhibitors [13,14]; TP53 status was determined by sequencing without p53 immunohistochemistry; histology was not centrally reviewed; public survival analyses were not adjusted for clinical covariates; and several subgroups were small.

In conclusion, TP53 PV was associated with worse OS in advanced EC and CCOC, and in HGSOC only with co-occurring KRAS PV; RB1 PV and, more modestly, MYC amplification were associated with worse OS in both organs, whereas KRAS, CTNNB1 and ARID1A associations were organ-specific. These findings may inform prognostic stratification and clinical trial design in gynecologic cancers.

## Author contributions

MP and LH conceived, designed and supervised the study. MP wrote the manuscript. PT and EC provided administrative support. CJ, NA, ST, PT, AVS and EC provided study materials. JS, SK, CJ, JZ, YK, HL, NA, PT, AVS and EC collected and assembled the data. MP, CJ, JS, SK, LH, HI and TG analyzed and interpreted the data. MP, LH, HI and TG revised the manuscript. All authors approved the final manuscript.

## Data availability

The KPNC Institutional Review Board has not approved placing individual-level StrataNGS data in a public repository. Researchers may request access through the KPNC Division of Research Data Sharing Workgroup. Public data are available through cBioPortal.

