## Supplementary Tables and Figures for "TP53 and Co-Occurring Pathogenic Variants Are Differentially Associated with Prognosis of Advanced Ovarian and Endometrial Carcinomas"

**Supplementary Table S1.** Multivariable Cox model of OS including all twelve PVs in ovarian carcinoma (analysis dataset, n = 1025; CCOC, MOC and serous histologies).

| Variable | HR | 95% CI | p |
| --- | --- | --- | --- |
| Age (per year) | 1.00 | 0.99-1.01 | 0.630 |
| Race: Asian vs White | 0.96 | 0.75-1.23 | 0.741 |
| Race: Black vs White | 1.08 | 0.73-1.60 | 0.700 |
| Race: Hispanic vs White | 0.96 | 0.68-1.37 | 0.837 |
| Race: Other vs White | 1.06 | 0.79-1.43 | 0.682 |
| CCI (per point) | 1.02 | 0.98-1.06 | 0.328 |
| ECOG PS 2-4 vs 0-1 | 2.01 | 1.54-2.63 | <0.001 |
| ECOG PS unknown vs 0-1 | 1.29 | 0.99-1.67 | 0.058 |
| Chemotherapy (yes vs no) | 0.95 | 0.78-1.17 | 0.648 |
| Mucinous vs clear cell | 2.07 | 1.08-4.00 | 0.030 |
| Serous vs clear cell | 0.95 | 0.61-1.49 | 0.827 |
| TP53 PV | 1.14 | 0.89-1.47 | 0.307 |
| BRCA1 PV | 0.66 | 0.48-0.91 | 0.012 |
| BRCA2 PV | 0.55 | 0.35-0.87 | 0.010 |
| KRAS PV | 0.71 | 0.52-0.98 | 0.039 |
| MYC amplification | 1.36 | 1.00-1.87 | 0.052 |
| PIK3CA PV | 1.02 | 0.74-1.42 | 0.891 |
| CDKN2A PV | 1.38 | 0.90-2.10 | 0.140 |
| CCNE1 amplification | 0.65 | 0.38-1.12 | 0.124 |
| RB1 PV | 1.79 | 1.13-2.84 | 0.013 |
| NF1 PV | 1.70 | 0.97-2.97 | 0.064 |
| PTEN PV | 1.29 | 0.77-2.18 | 0.339 |
| BRAF PV | 1.02 | 0.53-1.96 | 0.949 |
| ARID1A PV | 1.05 | 0.63-1.77 | 0.844 |

**Supplementary Table S2.** Multivariable HRs for OS of TP53 PV versus WT within molecular sub-cohorts of ovarian carcinoma (analysis dataset).

| Sub-cohort | TP53 PV vs WT, HR (95% CI) | p | Other PVs in model |
| --- | --- | --- | --- |
| KRAS PV | 2.13 (1.13-3.99) | 0.019 | BRCA, PIK3CA, MYC, ARID1A |
| KRAS WT | 0.99 (0.77-1.27) | 0.911 | BRCA, PIK3CA, MYC, ARID1A |
| BRCA1/2 PV | 0.69 (0.32-1.51) | 0.353 | KRAS, PIK3CA, MYC, ARID1A |
| PIK3CA PV | 0.80 (0.39-1.63) | 0.537 | BRCA, KRAS, MYC, ARID1A |

All models adjusted for age, race/ethnicity, CCI, PS and chemotherapy.

**Supplementary Table S3.** Characteristics of HGSOC by BRCA1/2 status.

| Characteristic | BRCA1 PV (n = 97) | BRCA2 PV (n = 50) | BRCA1/2 WT (n = 775) | p |
| --- | --- | --- | --- | --- |
| Age, median (range), years | 56 (32-84) | 64 (41-85) | 65 (26-94) | <0.001 |
| Asian race | 21 (21.6) | 10 (20.0) | 124 (16.0) | 0.247 |
| PS 0-1 | 77 (79.4) | 32 (64.0) | 577 (74.5) | 0.155 |
| PARPi received | 85 (87.6) | 38 (76.0) | 224 (28.9) | <0.001 |
| TP53 PV | 90 (92.8) | 40 (80.0) | 630 (81.3) | 0.037 |
| KRAS PV | 0 | 2 (4.0) | 72 (9.3) | 0.004 |
| MYC amplification | 12 (12.4) | 0 | 47 (6.1) | 0.009 |
| RB1 PV | 6 (6.2) | 1 (2.0) | 21 (2.7) | 0.155 |
| NF1 PV | 1 (1.0) | 5 (10.0) | 21 (2.7) | 0.006 |
| CCNE1 amplification | 1 (1.0) | 0 | 29 (3.7) | 0.150 |

Values are No. (%) unless noted. [Reconcile: BRCA1 PV n = 97 here versus 99 in Table 1.]

**Supplementary Table S4.** Concordance between the KPNC and public cBioPortal cohorts.

| Measure | KPNC | Public cBioPortal | Concordant |
| --- | --- | --- | --- |
| *Prevalence* |  |  |  |
| TP53 PV, HGSOC | 82.5% | 95.1% | No |
| TP53 PV, CCOC | 13.6% | 11.2% | Yes |
| TP53 PV, MOC | 64.3% | 74.5% | Yes |
| TP53 PV, endometrioid EC | 31.1% | 19.0%^d^ | No |
| TP53 PV, serous EC | 87.9% | 95.2% | Partly |
| KRAS PV, HGSOC | 8.0% | 1.4% | No |
| KRAS PV, CCOC | 24.2% | 13.0% | Partly |
| KRAS PV, EOC | 31.0% | 33.3% | Yes |
| KRAS PV, MOC | 82.1% | 72.7% | Yes |
| KRAS PV, endometrioid EC | 24.9% | 25.6% | Yes |
| KRAS PV, serous EC | 10.7% | 4.7% | Partly |
| MYC amplification, HGSOC | 6.4% | 13.6% | No |
| *OS, HR (95% CI)* |  |  |  |
| TP53 PV, ovarian | 0.95 (0.72-1.25)^b^ | 1.28 (1.00-1.62)^a,c^ | Direction differs |
| TP53 PV, endometrial | 1.61 (1.23-2.10)^c^ | 1.69 (1.36-2.10)^a,c^ | Yes |
| KRAS PV, ovarian | 0.72 (0.50-1.02)^b^ | 0.70 (0.43-1.14)^a,c^ | Yes |
| KRAS PV, endometrial | 0.90 (0.67-1.22) | 1.24 (0.95-1.62)^a,c^ | Yes (null) |
| TP53 × KRAS interaction, ovarian | 2.06 (0.98-4.35)^b^ | 1.87 (0.97-3.62) | Yes |
| TP53 × KRAS interaction, endometrial | NA | 0.63 (0.42-0.95) | NA |
| MYC amplification, HGSOC | 1.43 (1.05-1.95)^b^ | 1.08 (0.83-1.41) | Direction only |

^a^Stratified by histologic subtype. ^b^HGSOC. ^c^Altered without the other alteration of the TP53/KRAS pair versus neither. ^d^Public cohort includes early-stage disease. Public estimates are unadjusted for clinical covariates. Public TP53 alteration includes protein-altering mutation or deep deletion. NA, not available.

**Supplementary Table S5.** Exploratory analysis of TP53 variant class: hotspot gain-of-function (GOF; R175H, R248Q, R248W, R249S, R273H, R273L, R282W) versus other TP53 PVs.

| Cohort | GOF share of TP53 PVs | OS, GOF vs other TP53 PV, HR (95% CI) |
| --- | --- | --- |
| KPNC ovarian (HGSOC) | 103/762 (13.5%) | 1.06 (0.80-1.38)^a^ |
| KPNC endometrial | 64/314 (20.4%) | 1.12 (0.81-1.55)^b^ |
| Public ovarian | 14.6% | 1.09 (0.89-1.34)^c^ |
| Public endometrial | 24.4% | 1.03 (0.83-1.27)^c^ |
| Pooled, four cohorts |  | 1.07 (0.95-1.21); I² 0% |

^a^Multivariable HGSOC model. ^b^Adjusted for age and PS; versus WT, HR 1.80 (1.28-2.54) for GOF and 1.62 (1.27-2.06) for other TP53 PVs. ^c^Stratified by histologic subtype. Pooled estimate is inverse-variance fixed-effect.

### Supplementary Figures

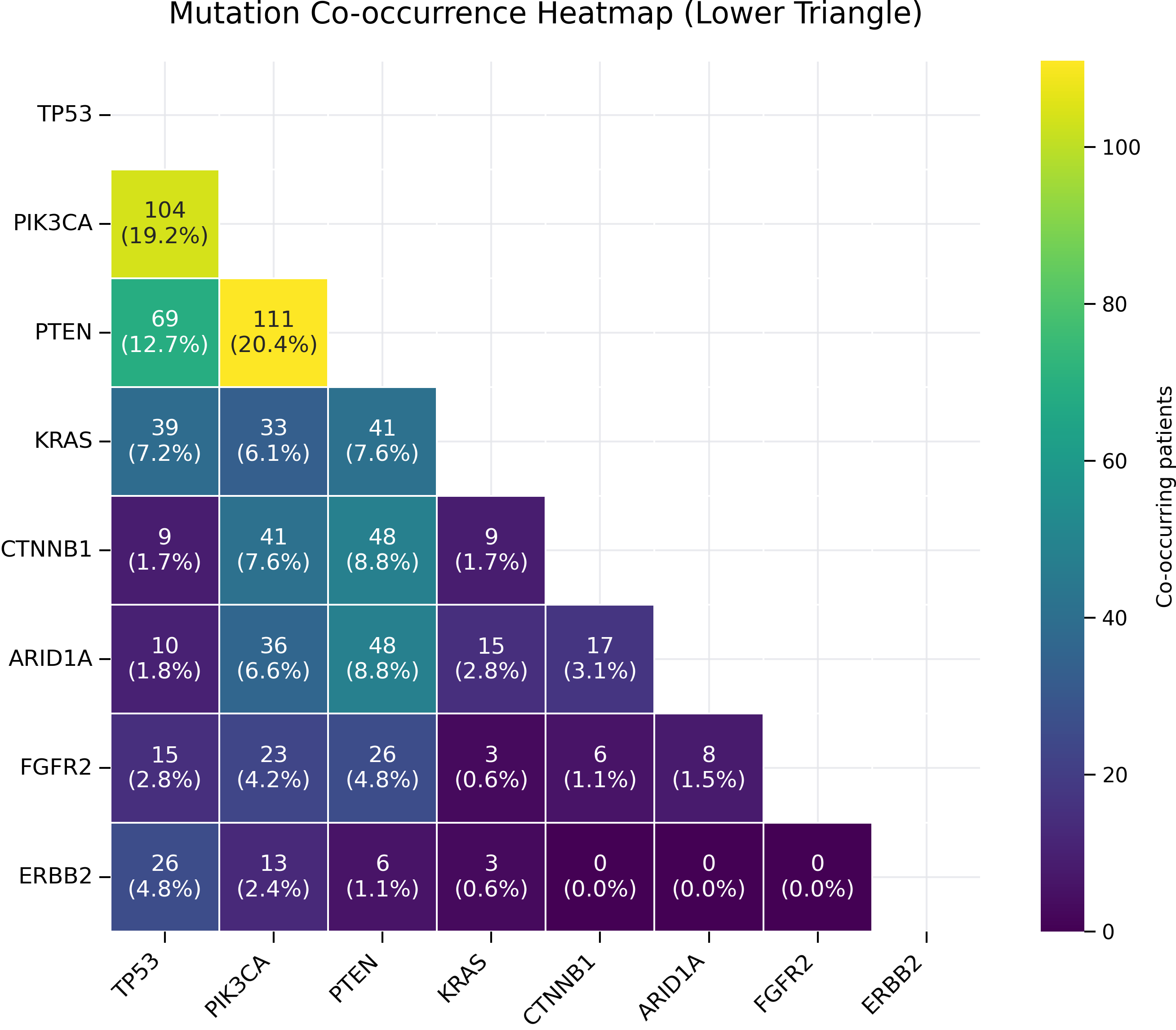

**Supplementary Fig. S1.** Co-occurrence of the most common PVs in endometrial carcinoma.

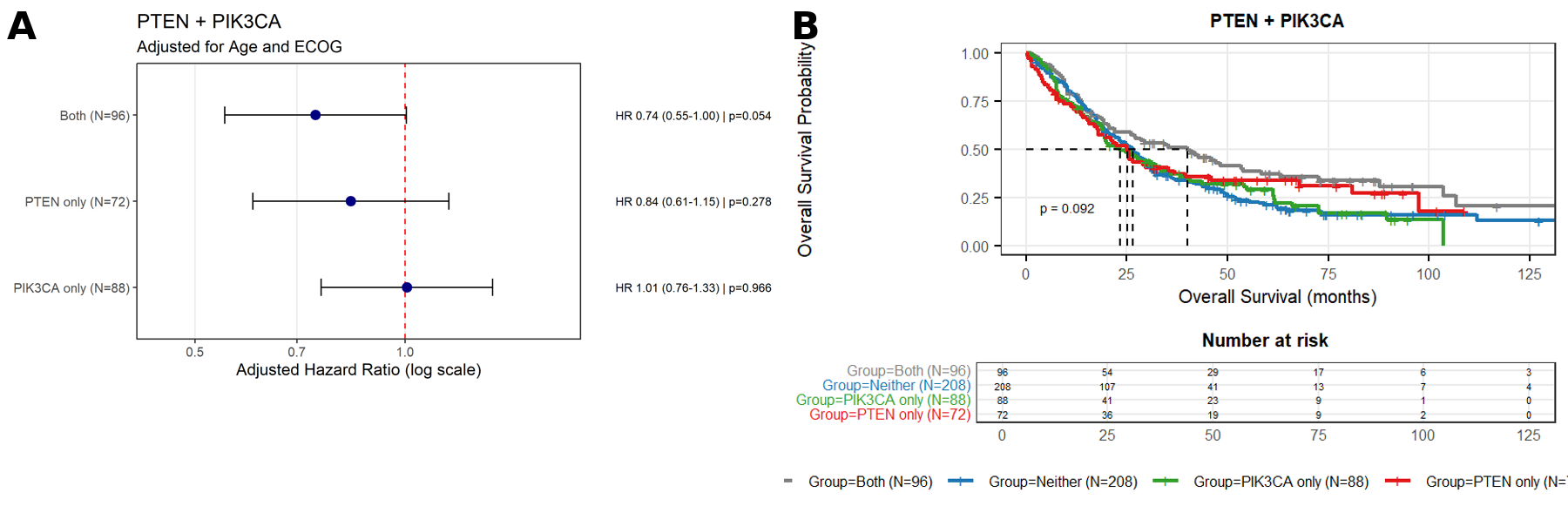

**Supplementary Fig. S2.** (A) HRs (adjusted for age and PS) and (B) Kaplan-Meier OS for PTEN and PIK3CA co-alteration groups in endometrial carcinoma (p = 0.092).

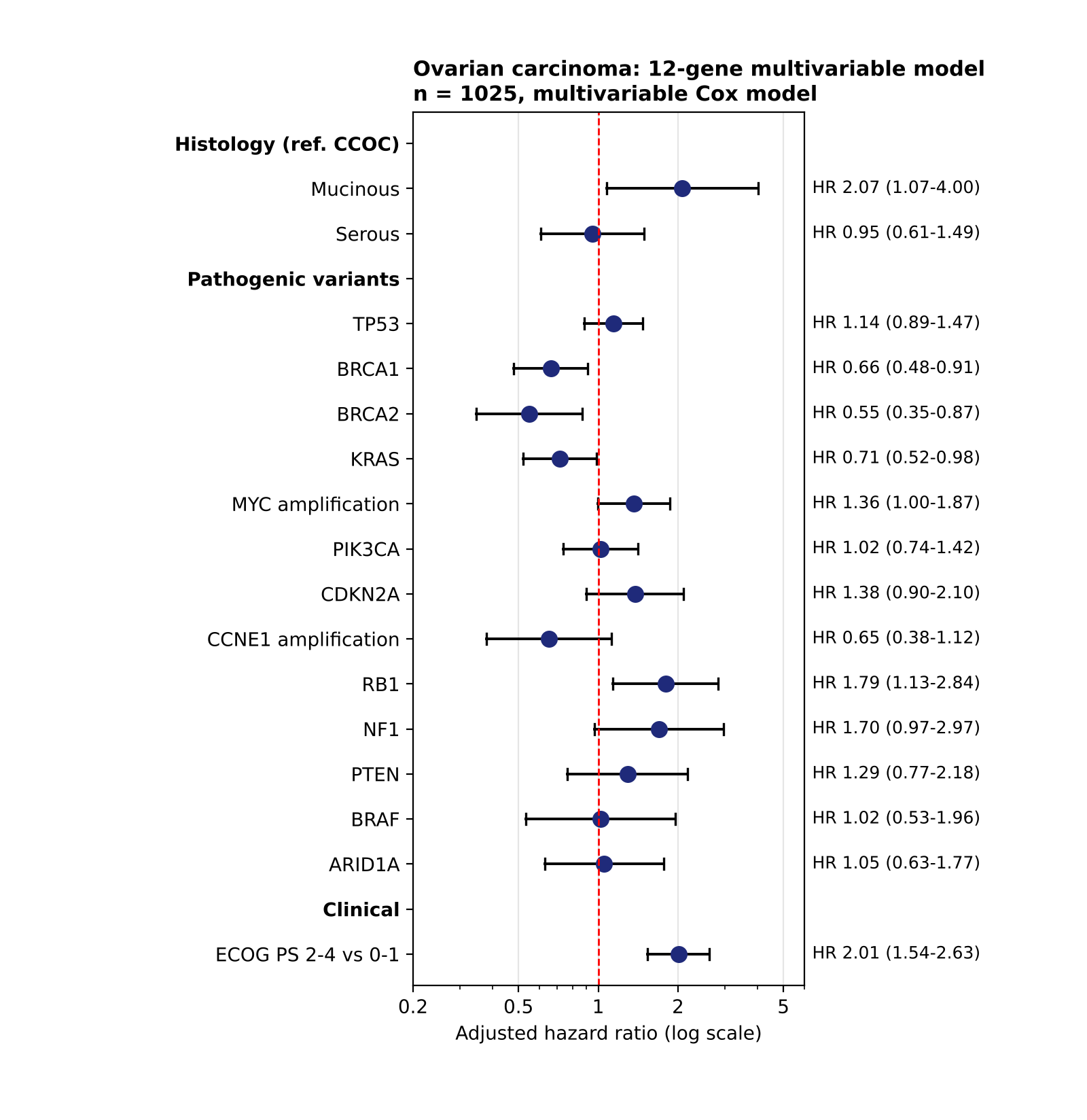

**Supplementary Fig. S3.** Twelve-PV multivariable Cox model in the ovarian analysis dataset (Supplementary Table S1).

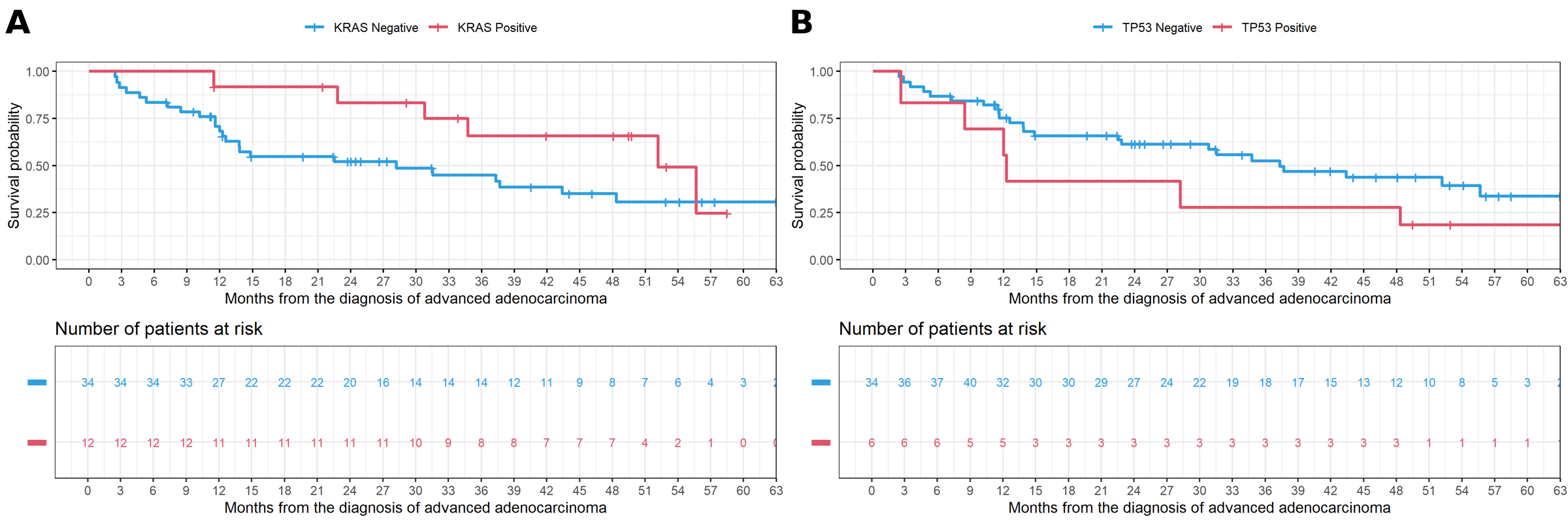

**Supplementary Fig. S4.** Kaplan-Meier OS of CCOC by (A) KRAS PV status (p = 0.25) and (B) TP53 PV status (p = 0.17).

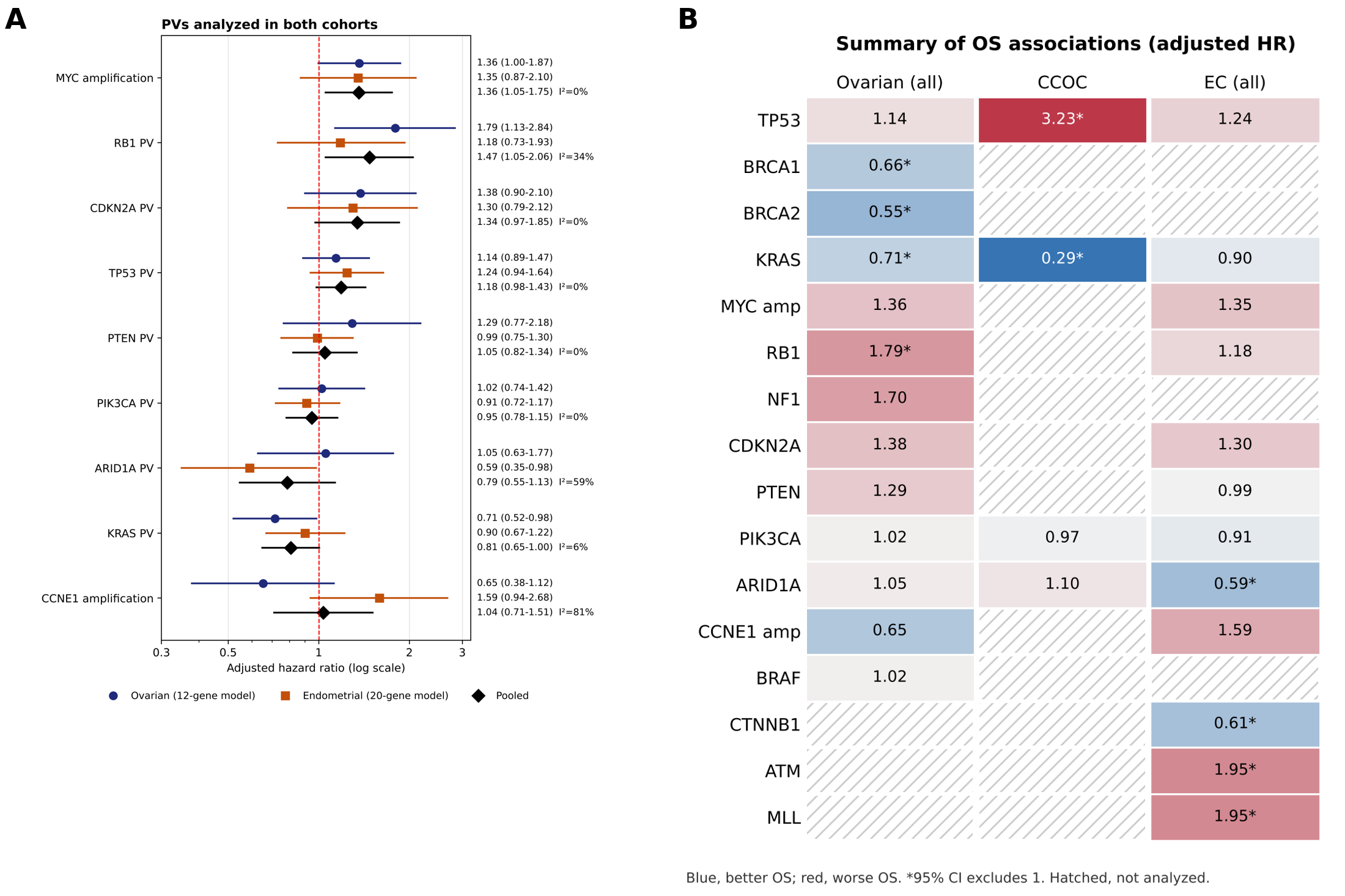

**Supplementary Fig. S5.** Shared and organ-specific associations of PVs with OS. (A) Adjusted HRs in ovarian carcinoma (twelve-PV model) and EC (twenty-PV model) with fixed-effect pooled estimates and I². (B) Summary of adjusted HRs in all ovarian carcinoma, CCOC and EC; blue, better OS; red, worse OS; *95% CI excludes 1; hatched, not analyzed.

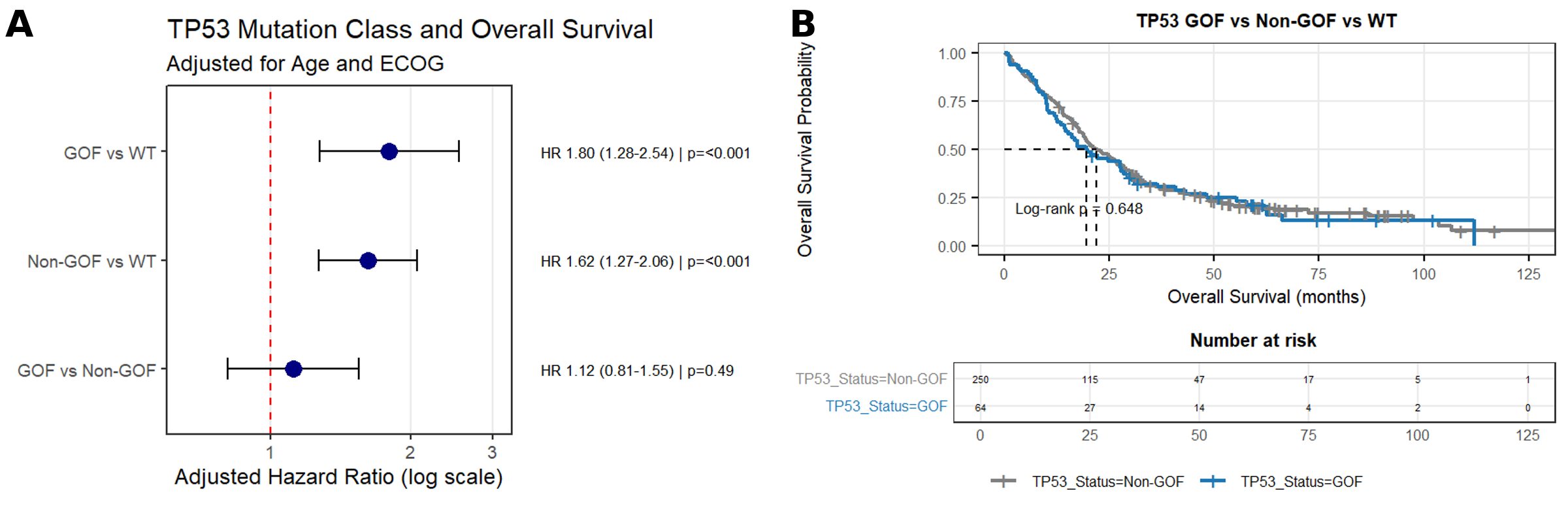

**Supplementary Fig. S6.** TP53 variant class and OS in endometrial carcinoma. (A) HRs for hotspot GOF versus WT, other TP53 PV versus WT, and GOF versus other TP53 PV, adjusted for age and PS. (B) Kaplan-Meier OS of GOF (n = 64) versus other TP53 PVs (n = 250) (log-rank p = 0.648).
